# Data-driven identification of previously unrecognized communities with alarming levels of tuberculosis infection in the Democratic Republic of Congo

**DOI:** 10.1101/2021.12.09.21267511

**Authors:** Mauro Faccin, Olivier Rusumba, Alfred Ushindi, Mireille Riziki, Tresor Habiragi, Fairouz Boutachkourt, Emmanuel André

## Abstract

When access to diagnosis and treatment of tuberculosis is disrupted by poverty or unequal access to health services, marginalized communities not only endorse the burden of preventable deaths, but also suffer from the dramatic consequences of a disease which impacts one’s ability to access education and minimal financial incomes. Unfortunately, these pockets are often left unrecognized in the flow of data collected National tuberculosis reports, as localized hotspots are diluted in aggregated reports focusing on notified cases. Such system is therefore profoundly inadequate for identifying these marginalized groups, which urgently require adapted interventions.

We computed an estimated incidence-rate map for the South-Kivu province of the Democratic Republic of Congo, a province of 6.3 million inhabitants, leveraging available data including notified incidence, level of access to health care and exposition to identifiable risk factors. These estimations were validated in a prospective multi-centric study.

We could demonstrate that combining different sources of openly-available data allows to precisely identify pockets of the population which endorses the biggest part of the burden of disease. We could precisely identify areas with a predicted annual incidence of *>* 1%, a value three times higher than the national estimates. While hosting only 2.5% of the total population, we estimated that these areas were responsible for 23.5% of the actual tuberculosis cases of the province. The bacteriological results obtained from systematic screenings strongly correlated with the estimated incidence (r=0.86), and much less with the incidence reported from epidemiological reports (r=0.77), highlighting the inadequacy of these reports when used alone to guide disease control programs.

## Introduction

Worldwide, it is estimated that up to 4 million patients affected by active tuberculosis disease (TB) are left untreated every year. In Africa, up to 50% of patients requiring care are left undiagnosed today, and while drug-resistant TB remains a major concern, there are probably more transmission and deaths related to TB under-detection than due to drug resistance^1^.

The direct and indirect costs supported by these millions of people in need of care are both the cause and the consequence of the socio-economic impact of TB on the poorest populations. In this sense, inadequate access to care is a major contributor to the dramatic spiral of poverty and uncontrolled disease transmission.

Recognizing the importance of this problem, the World Health Organization (WHO) recommends to perform systematic TB screening in high-risk communities, and since a decade, numerous “active case finding” (ACF) pilot projects have been supported and implemented in high burden countries. The very irregular level of efficacy of these programs is intrinsically related to the difficulty to identify pockets of the population with the highest burden of disease. Performing systematic screening in a population with low incidence or having a facilitated access to health services outside screening campaigns will lead to very limited additional cases found. Additionally, it will typically impact staff motivation and require difficultly scalable human and financial resources^1^,2.

Pushed by the necessity to achieve a significant yield, a typical reccomandation is to perform systematic screening in households of TB patients and among people living with HIV. Although these indications are well proven to be useful, restricting systematic screening to these very limited groups will structurally miss the opportunity to detect the majority of TB cases, in particular in the context of structural under-diagnosis of TB and HIV such as experienced in the DRC. It has further been described that within households of active TB cases, the source of new infections is very often different from the presumed index case^3^. These oservations illustrates the need to extend systematic screening beyond the current recommended perimeter, in particular in areas with very high incidence of the disease^4^.

These recommendations are in practice inapplicable, as countries lack tools to identify pockets of the population where high levels of under-notification hides dramatic incidence rates of TB. We develop a tool which would precisely identify the pockets of the population were the majority of TB cases are undetected. In this context, such tool would allow guiding highly efficient ACF interventions, and would allow avoiding over- or under-utilization of community workers and medical resources.

## Results

We introduce a new approach to ACF planning which is in two-folds: it combines a data-driven detection of high-incidence pockets of the disease and a digital assessment of the individual risk as triage.

Firstly, we collect openly-available datasets that describe environment characteristics such as the population density, the presence of the local health care system or the closeness to mining sites. A data-driven prediction algorithm combines these datasets with the information from the local TB reports to precisely identify localized pockets of very high circulation of TB which can be defined as an incidence rate above 1000/100000 (1%). A representation of these calculations on a map of the South-Kivu province of the Democratic Republic of Congo (DRC) is illustrated in Figure 1. In this map, colour codes represent the estimated incidence rate for active pulmonary tuberculosis. This map illustrates the significant variations in the predicted incidence within the province: while the vast majority of the surface of the province shows predicted incidence rates below 0.1%, only very limited areas actually show a risk of above 1%. In South-Kivu, the share of population living in high-risk zones, with a predicted incidence rate above 1%, is only about 2.5%. This same population is expected to host more than 20% of active TB cases.

**Figure 1.**
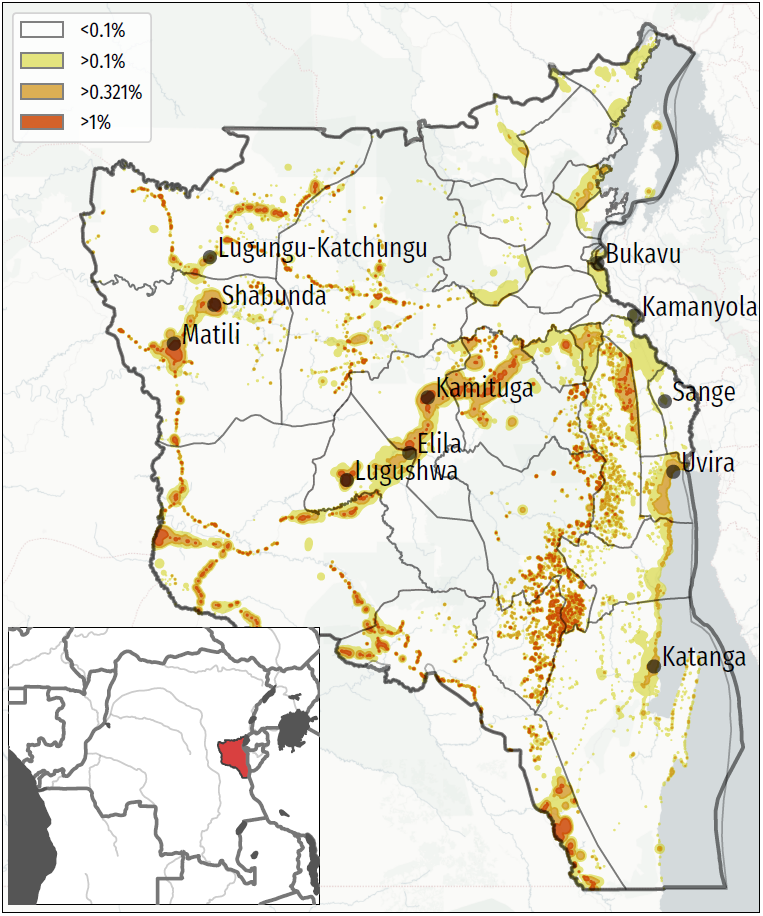
Mapping of TB predicted incidence rate for the South-Kivu province of the Democratic Republic of Congo (inlay). No color: below 0.1%; Yellow : between 0.1% and national average (0.322%); Orange : between 0.322% and 1%; Red : above 1%.

Secondly, in the communities highlighted by the estimated incidence rate an ACF intervention are performed with the aid of a digitally supported questionnaire and the MediScout© application stack as triage.

### Study setting and study design

We performed a multi-centric prospective study in the South-Kivu province of the DRC, a region at the border with Rwanda and Burundi and facing a high burden of TB, partly due to a situation of over 20 years of conflicts and population displacements. As most eastern provinces of DRC, South-Kivu has important artisanal and industrial mining sectors. The South-Kivu province hosts a population of 5,8 million inhabitants, and is divided in 34 health zones. In total, 113 health facilities providing basic TB diagnostic and treatment services.

Two main elements were evaluated in this study: the community risk measure represented by the map of predicted incidence rate and the individual risk measure represented by the questionnaire and digital tool.

Firstly, we aimed to evaluate the ability of the relative incidence rate prediction map to correctly predict the incidence rate of TB in particular communities. The major outcome is the ability to segregate communities with a very high rate of TB (>1%) from other communities not systematically eligible for ACF interventions as per the WHO criteria.

To do so, we included 11 urban, semi-urban and rural communities distributed in 11 health zones of the province. The choice of these communities was made in order to cover a wide range of TB predicted incidence, ranging from 0.1% to 2.3% (see Table 1). Some zones at high predicted risk such as Itombwe and Minembwe had to be excluded due to ongoing insecurity issues. We included in this study remote communities such as Matili and Lugushwa which were only accessible using local planes combined by two days of travel on a motorcycle. Table 2 provides detailed information on the notified and predicted incidence for each of these communities. In this table, we observe sensible differences between the notified incidence rate and the predicted incidence rate, the predicted incidence rate being up to 10 times higher than the notified incidence rate in the same area. An example of this situation is the city of Shabunda, a rural and mining area with a very low coverage of health structures.

**Table 1.**
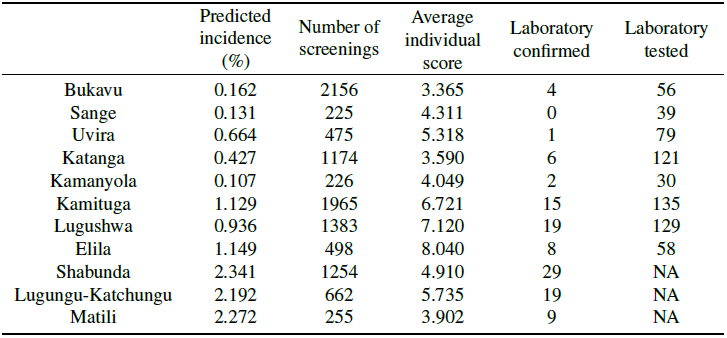
Project statistics disaggregated by location.

|  | Predicted incidence (%) | Number of screenings | Average individual score | Laboratory confirmed | Laboratory tested |
| --- | --- | --- | --- | --- | --- |
| Bukavu | 0.162 | 2156 | 3.365 | 4 | 56 |
| Sange | 0.131 | 225 | 4.311 | 0 | 39 |
| Uvira | 0.664 | 475 | 5.318 | 1 | 79 |
| Katanga | 0.427 | 1174 | 3.590 | 6 | 121 |
| Kamanyola | 0.107 | 226 | 4.049 | 2 | 30 |
| Kamituga | 1.129 | 1965 | 6.721 | 15 | 135 |
| Lugushwa | 0.936 | 1383 | 7.120 | 19 | 129 |
| Elila | 1.149 | 498 | 8.040 | 8 | 58 |
| Shabunda | 2.341 | 1254 | 4.910 | 29 | NA |
| Lugungu-Katchungu | 2.192 | 662 | 5.735 | 19 | NA |
| Matili | 2.272 | 255 | 3.902 | 9 | NA |

**Table 2.** Distribution of reported and estimated incidence on the communities interested in this work.

| Communities<br>(Health zone) | Reported<br>incidence<br>(2017) | Estimated<br>incidence | Current<br>detection rate<br>(estimated) |
| --- | --- | --- | --- |
| Kamanyola (Nyangezi) | 100 | 110 | 91% |
| Sange (Ruzizi) | 40 | 130 | 31% |
| Bukavu (Ibanda, Kadutu, Bagira) | 100 | 160 | 63% |
| Katanga (Fizi) | 150 | 430 | 35% |
| Uvira (Uvira) | 170 | 660 | 26% |
| Lugushwa (Kitutu) | 170 | 940 | 18% |
| Kamituga (Kamituga) | 240 | 1129 | 21% |
| Elila (Kitutu) | 260 | 1149 | 23% |
| Lugungu-Katchungu (Lulingu) | 250 | 192 | 11% |
| Matili (Shabunda) | 260 | 272 | 11% |
| Shabunda (Shabunda) | 170 | 341 | 7% |

Secondly, we aimed to evaluate the performance of the MediScout© mobile app, including the built-in individual risk assessment tool (see Figure 2). Therefore, we partnered with a local organization experienced in TB ACF and trained a project coordinator and 20 research assistants involved in community-outreach activities for the utilization of the MediScout© applications. This training included the initiation of ACF missions, individual screenings and referral of patients to local health clinics. These screeners, originating from the provincial capital city Bukavu, visited all study areas, where they were accompanied by local community health workers to facilitate the collaboration of the community.

**Figure 2.**
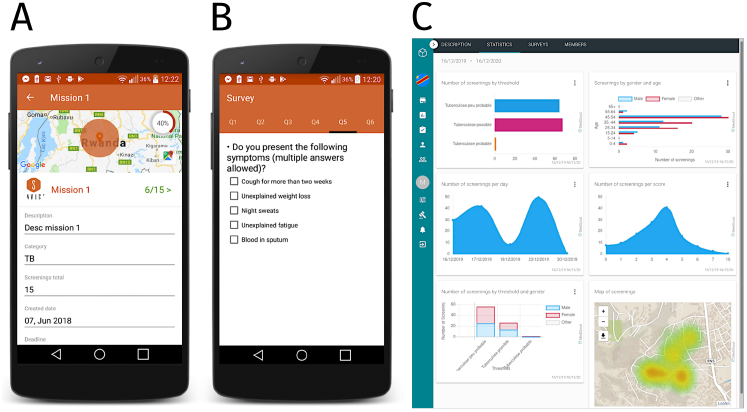
The Mediscout© mobile and web applications. (A) Mobile application with interface for mission acceptance by the community health-worker. (B) Questionnaire in the mobile application. (C) Statistics of the missions in the web application (note the mapping of the screenings at the bottom).

### Performance of the relative incidence rate prediction tool

We used the individual risk-assessment questionnaire to evaluate the performance of the incidence rate prediction algorithm. This questionnaire, based on a combination of TB-related symptoms, personal exposition to TB and personal history of TB, was considered as a good proxy for estimating the level of circulation of TB in a community (see 3). Overall, in the 11 study sites, 13841 people agreed to respond to the individual risk-assessment questionnaire. 8322 (60.1% of total) questionnaires originated from areas at low predicted incidence rate (*<* 1%) and 5519 (39.9%) questionnaires originated from areas at high predicted incidence rate (> 1%) (see Table 4). In areas with low predicted incidence, 55.5% of the responders had a low risk (score *<*4) and 44.5% had a high risk for TB (score *≥*4). In comparison, in areas with higher predicted incidence, 32.5% of the responders had a low risk (score *<* 4) and 67.5% had a high risk for TB (score *≥*4).

The individual risk score is comprised between 0 and 20, as is computed based on a diversity of questions including related to the presence of several TB-related symptoms and different elements reflecting the intensity of exposition to TB. We looked at the distribution of these scores in the high and low predicted incidence areas.

The median and the inter-quartile ranges of the risk score in low or high incidence predicted areas was of 3(2-6) and 5(3-9) respectively, highlighting the different, although partly overlapping, distribution of scores in the two populations (p-value=0). Figure 3 shows the distribution of the risk score in the two types of settings.

**Figure 3.**
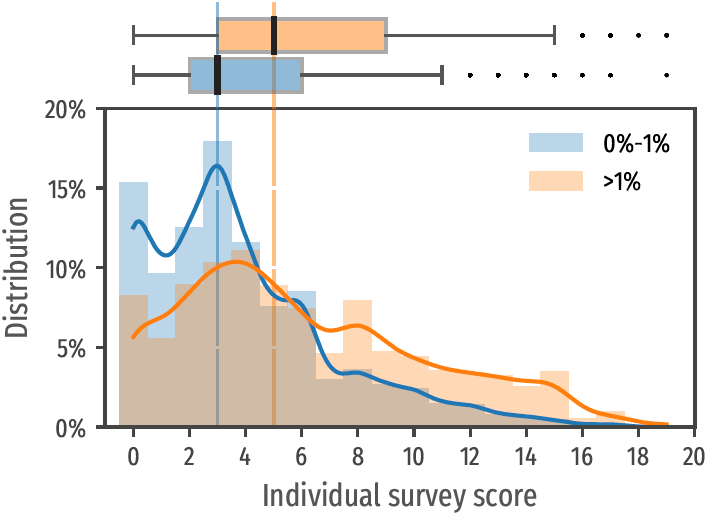
Distribution of individual risk for the population located in low risk zones (blue) where the predicted incidence rate is lower than 1% and for the population in high risk zones (orange). Note that the proportion of lower scores (<4) is higher in the lower risk communities (bottom). Score medians in both sub-populations (3 and 5 respectively) with quartiles are reported in the upper box plot.

Figure 3 (top) illustrates how the individual risk score in the two populations at high and low risk follows different distributions (with p-value=0). It also illustrates the high dispersion of risk scores within these areas, reflecting the heterogeneity of individual risk in each community: the individual score median as well as the confidence intervals increase in areas at higher risk.

### Performance of the individual-risk assessment questionnaire

We used Ziehl-Neelsen microscopy as a reference method to evaluate the performance of the individual-risk assessment questionnaire.

In total 1153 laboratory tests were performed. Screeners were trained to suggest a laboratory test only among people presenting a cough (regardless of the total score) or having an individual risk score *≥*4. Within the performed tests, 112 individuals were diagnosed with laboratory-confirmed pulmonary TB (9.7% positivity). Unfortunately, some laboratory outcomes in few remote facilities resulted unlinked from the corresponding questionnaire and were discarded from this analysis. The positivity rate was 8.8% (n=55/626) for those with a score *≥*4 and 12.3% for those with a score *≥*8 (n=48/389).

No positive laboratory results were reported among people with a score lower than 4, regardless of the presence of cough (21 lab tests). The proportion of positive smears increased together with the risk score as illustrated in Figure 4, supporting the effectiveness of the mobile questionnaire as triage.

**Figure 4.**
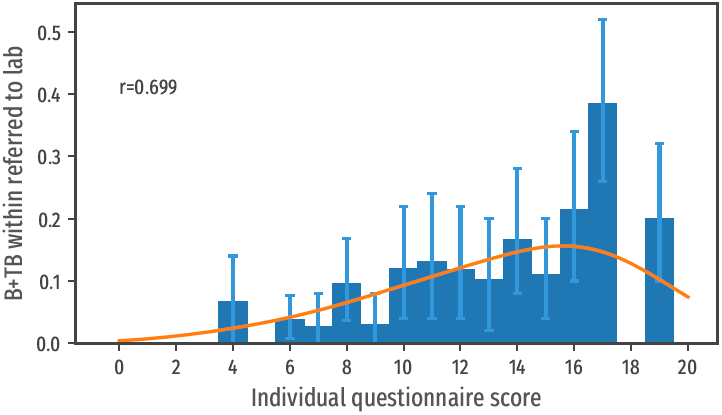
Proportion on laboratory confirmed TB cases per score class.

### Performance of the integrated prediction as a policy-decision tool for ACF disease control programs

To assess the performance of MediScout© as a technical tool that can be used by TB-program managers and community-outreach organizations to optimize the efficiency of ACF interventions, we compared the predicted incidence rate with the reported incidence of bacteriologically-confirmed pulmonary cases.

Although the populations originating from areas with high risk (> 1% predicted incidence) and areas with low risk were of similar size, of the 112 individuals diagnosed with bacteriologically-confirmed pulmonary TB, 91 (81.25%) originated from the predicted zones at very high risk and 21 (18.75%) originated from zones predicted to have an incidence rate *<*1%.

Figure 5 (left) shows a strong association (r=0,861) between the predicted incidence rate and the proportion of bacteriologically-confirmed pulmonary TB (B+TB) cases identified, with a nearly linear correlation between the predicted incidence and the observed yield of the ACF interventions (with a fitting line with a slope of 0.95). In comparison, Figure 5 (right) illustrates a much less performant correlation between the historically notified cases and the actual incidence observed in this study associated with the heterogeneous level of under detection of TB.

**Figure 5.**
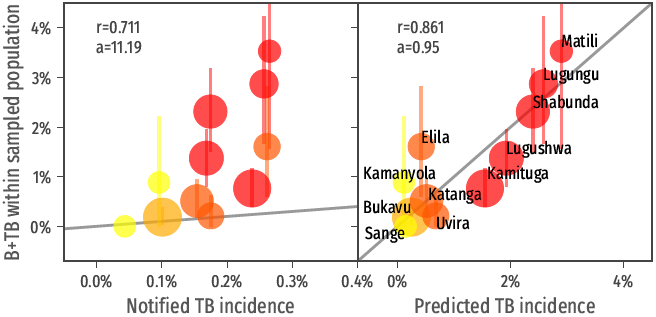
Correlation of the predicted incidence with the measured incidence within sampled population (left). Note the high value of the correlation coefficient *r*. The parameter *a* represent the slope of the fitting line. As a term of comparison, the correlation of the incidence extracted from the health system reports with the measured incidence within the sampled population (right). Note in this case the lower correlation coefficient and the slope of the fitting line (the measured incidence surpass 10x the incidence of reported cases).

## Discussion

One of the major limitations of this study is that the diagnosis of TB relied uniquely on series of three Ziehl-Neelsen microscopy tests per patient. This technique probably underestimated the level of active TB disease as it is known to have a limited sensitivity and would therefore miss a proportion of paucibacillary infections. The real incidence may thus have been higher than reported in this study. Despite this limitation, we believe that performing this evaluation in a real-life setting, which includes the technical limitations experienced by health workers, is probably more informative than a study which would create an artificial bias by including technologies which are currently inaccessible for the vast majority of the population.

Very interestingly, we could show that using data-driven predictions and setting a threshold at a predicted incidence of 1% allows prioritizing ACF interventions in well-circumcised pockets of the population where over 80% of the cases found through ACF reside. This illustrates that in such setting with uncontrolled transmission, the majority of the people experiencing active TB disease will not access the health system if they are not actively identified and supported by outreach interventions.

Another major outcome of this study is that the planning and prioritization of TB control interventions should not be made upon historical notification reports. In such cases, the decision process would contribute to under-estimate the actual level of disease transmission in the most vulnerable pockets of the population without or with limited access to the health system. This study highlights that these historical notifications reports should be integrated with demographical, geographical and social data in order to optimally inform public health authorities and funders about where to prioritize the implementation of complementary interventions.

## Methods

### Relative incidence rate prediction

We gathered openly available data linked to the risk of TB to compute a relative risk level for each precise location. The different data sources and their utilization in our model are described hereunder.

Population distribution and density information was extracted from the Worldpop project^5^,6, an initiative which provides an estimation of the population density with a resolution of approximately 100 meters. These estimations result from combined geospatial datasets with available aggregated count data^7^. Further, additional information on the location of urban and residential environments was gathered from the OpenStreetMap project.

We then combined the demographic data with the most granular level of epidemiological surveillance, being, in the context of DRC where this study took place, the Health Zone quarterly notification reports. These were used to make a baseline distribution of the TB cases. For this baseline distribution, we assumed areas with low population density have a lower TB incidence-rate compared to higher density areas. Locations with a population density lower than 10 people per square kilometre were ignored in this distribution model.

We further used the location and type of health facilities extracted from the Global Healthsites Mapping Project^8^ in collaboration with Openstreetmap^9^. We used the distance between each community and the nearest health facility to estimate the phenomenon of under-detection of TB, which is correlated to the difficulties for each community to access health services, assuming that “far” from health facilities, 50% of the real TB cases are missed. Furthermore, the type of health facility local clinical versus hospitals was also used in the risk estimation.

Finally, silica exposure, in particular when linked to mining activities, is a risk factor for TB. Since the South-Kivu province, where this study took place, has an important mining activity, we integrated in the risk computation the location and size of mines, accessible through the IPIS Research project^10^ and Openstreetmap^9^. The predicted risk of TB was correlated with the presence of mines in the same geographical areas. We assumed that in mining environments, the incidence rate of TB cannot be lower than of 0.5% or 500/100000.

### Individual risk prediction

The main output that was to be achieved by the individual assessment questionnaire was to be able to yield a positivity rate greater than 10% among people identified as being at high risk for active TB.

To build this individual assessment tool, we took into consideration previous observations highlighting that individual risk for TB infection can be extrapolated from a combination of elements including symptoms and the type of exposition to TB^11^.

Our questionnaire for evaluating the individual risk for TB was therefore based on the presence or absence of several TB-related symptoms and the precise context of an eventual exposition to TB. The elements captured and the weight given to each element is described in Table 3. The survey generates a total score in the range 0-20. In this study, all people with a score higher than 4 and those with a cough (regardless of the total score) were referred for a laboratory test.

**Table 3.** Elements and corresponding weights used to evaluate individual risk for TB in the questionnaire.

| Item evaluated during questionnaire | Score |  |
| --- | --- | --- |
| Unexplained fatigue | 1 | **Symptoms**<br>**(max score = 10)** |
| Unexplained weight loss | 1 |  |
| Night sweats | 1 |  |
| Temperature above 38°C | 1 |  |
| Cough (< 15 days) | 1 |  |
| Cough (> 15 days) | 2 |  |
| Blood in sputum | 2 |  |
| Chest pain | 1 |  |
| Close and persistent contact | 2 | **Individual TB exposure**<br>**(max score = 8)** |
| Occasional close contacts | 1 |  |
| Contacts in poorly ventilated area | 1 |  |
| Contacts occurred less than 2 years ago | 1 |  |
| Living or lived in military camp | 1 |  |
| Living or lived in mining camp | 1 |  |
| Living or lived in prison | 1 |  |
| History of abandoned treatment | 1 | **Individual TB history**<br>**(max score = 2)** |
| History of failed treatment | 1 |  |

**Table 4.** Participant distribution over high and low predicted incidence areas.

| Estimated<br>incidence rate | Participants<br>with score < 4 | Participants<br>with score ≥ 4 | Total number<br>of participants |
| --- | --- | --- | --- |
| < 1% | 55.5% (4614) | 44.5% (3708) | 8322 |
| ≥ 1% | 32.5% (1792) | 67.5% (3727) | 5519 |

### MediScout© solution

The MediScout© web application (developed by Savics, Belgium) was used as an integrated end-user interface allowing to visualize the incidence prediction map and plan geolocalized ACF campaigns. The MediScout© mobile application (Savics, Belgium) was used to guide community-based health workers in the restricted area of interest, while receiving from the remote web application ACF missions to perform. Further, the app was used to go through the individual questionnaires when in direct contact with the study participants, to automatically compute the individual risk and send the results of these questionnaires back to the remote server for later analysis, see Figure 2 for a visualization (Figure 2.A: mobile application mission interface, Figure 2.B mobile application questionnaire interface). The MediScout© mobile app allows uploading information to the MediScout© web interface when internet or 3G connectivity were available. This system allows full traceability of all community-based screening events, including GPS location, demographics of the population covered and individual scores.

Based on these reports, aggregated statistics and geographic locations are reported automatically and in real-time (Figure 2.C: web app interface with example of data visualization tools and intervention mapping at the bottom).

### Cases distribution from population density

The cases reported by the local health system are disaggregated according to the local population density inspired by the equilibrium solution of a simple compartmental model (SIS) of an endemic infected population^12^:

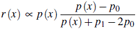

where *p*_0_ represents the minimal value of the population density for which the endemic infected population survive. Furthermore, *p*_1_ accounts for the relation between the population density and the transmission parameter of the disease. A normalization is applied in order to recover the same aggregated figures. We choose *p*_0_ = 10 inhabitants per km^2^ and *p*_1_ = 1000 inhabitants per km^2^.

### Choice of individual risk threshold

In the present prospective study, the community health workers suggested single individuals to refer to a laboratory for a microscopy test if their individual-risk score reached at least the conservative threshold value of 4. The choice of such conservative value is dictated by the specific testing aims of the current study. We computed the sensitivity and specificity of the survey based on different thresholds (see Figure 6). The chosen threshold presents a sensitivity=1.0 (no screening with individual score below 4 where confirmed positive to TB in laboratory) and a specificity=0.46. The choice of a threshold value equal to 6 would still have high sensitivity=0.98 while increasing the specificity=0.67 (and decreasing the number needed to test in laboratory to find a positive case). Applying a threshold at 6 would have saved 2692 (36%) tests, but only 1 case (^*∼*^ 0%) would have been missed.

**Figure 6.**
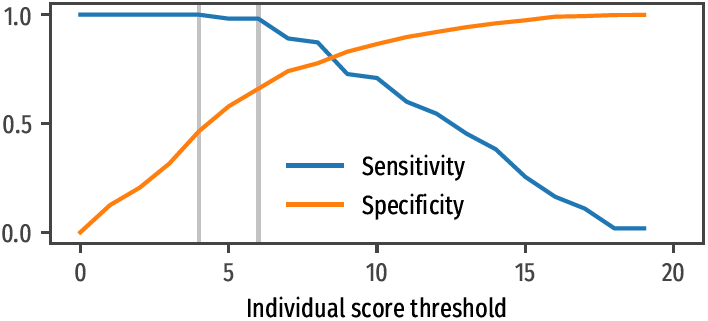
Expected sensitivity and specificity if the threshold to refer subjects to lab is changed to a different value. The vertical lines correspond to a threshold of 4 and 6. A threshold of 4 is maybe too conservative; in other contests, one can safely use 6.

## Data Availability

All data produced are available online at https://maurofaccin.github.io/cartotb

## Acknowledgements

MF was partially funded by Innoviris grant number D1.31402.007. MF’s current affiliation is Institut de Recherche pour le Développement (IRD) and Centre Population et Développement (CEPED), Université de Paris, France.

## Author contributions statement

**MF** Study design, data analysis, writing;

**OR** Study design, data collection;

**AU, MR, TH** Data collection;

**FB** Data collection, data analysis;

**EA** Study design; conceptualization; formal analysis; writing.

## Additional information

- The authors declare that they have no competing interests.
- The corresponding author is responsible for submitting a competing interests statement on behalf of all authors.
- The study and its protocols have been approved by the *Comité Institutionnel d’Ethique de la Santé* of the *Université Catholique de Bukavu* with reference number UCB/CIES/NC/07/2019.
- All subjects participating in this study (or their legal guardians) gave its informed consent. All methods were carried out in accordance with the relevant guidelines and regulations.

## Notes

### Competing Interest Statement

The authors have declared no competing interest.

### Funding Statement

This study was funded by Innoviris (Belgium)

### Author Declarations

The study and its protocols have been approved by the Comité Institutionnel d'Ethique de la Santé of the Université Catholique de Bukavu with reference number UCB/CIES/NC/07/2019.

